# Implementing relational continuity in general practice - understanding who needs it, when, to what extent, how and why: A realist review protocol

**DOI:** 10.1101/2025.04.15.25325890

**Authors:** Victoria Tzortziou Brown, Sophie Park, Kamal Mahtani, Stephanie Taylor, Emily Owen-Boukra, Jonathan Taylor, Owen Richards, Sultana Begum, Geoff Wong

**Affiliations:** Wolfson Institute of Population Health, Queen Mary University of London, UK; Nuffield Department of Primary Care Health Sciences, University of Oxford, UK

## Abstract

**Introduction:** Relational continuity of care (RCC) refers to the sustained therapeutic relationship between a patient and a clinician, which fosters trust, enhances communication, and facilitates the accumulation of knowledge about the patient. RCC is associated with enhanced patient outcomes, reduced hospital admissions, lower mortality rates, decreased healthcare costs, and improved patient experience. Despite these benefits, reorganisations within the NHS and workforce challenges have led to an increased reliance on multidisciplinary and part-time working, resulting in fragmented care and a decline in RCC. Our study aims to explore who needs RCC, under what circumstances, to what extent, and why, with the goal of informing optimal implementation strategies.

**Methods and Analysis:** We will conduct a realist review to develop an evidence-based programme theory explaining the mechanisms underlying RCC, the populations that benefit most, the contextual factors influencing RCC, and effective care models. Following Pawson’s five iterative stages, we will: (1) locate existing theories, (2) search for relevant evidence, (3) select appropriate articles, (4) extract and organise data, and (5) synthesise findings to draw conclusions. A stakeholder advisory group, comprising policymakers, healthcare professionals, public contributors, and patients, will be engaged throughout the process. We will adhere to Realist and Meta-narrative Evidence Synthesis: Evolving Standards (RAMESES) quality standards for realist reviews to ensure methodological rigour.

**Dissemination and ethics:** Our findings will inform practical, evidence-based recommendations for optimising RCC within general practice. Outputs will include peer-reviewed publications, conference presentations, plain English summaries, social media infographics, a short video, and end-of-study events. Collaborations with stakeholders and public involvement will ensure both accessibility and impact. Ethical review is not required for this evidence synthesis.

## Introduction

Relational continuity of care (RCC) refers to the sustained therapeutic relationship between a patient and healthcare professionals, which fosters trust, communication, and clinical responsibility. ^1^RCC in general practice is linked to improved patient outcomes, reduced hospital admissions, lower mortality rates, decreased healthcare costs, and enhanced patient experience. ^2, 3, 4^ Despite these benefits, there is little consensus on how to deliver RCC effectively and for whom it should be prioritised.

NHS reorganisations, workforce shortages, and the expansion of multidisciplinary teams have contributed to a fragmented healthcare system, resulting in reduced RCC. Policies that prioritise timely access over continuity have further eroded RCC, with only 16% of patients in 2023 consistently able to see their preferred GP. ^5^ Calls for incentivising RCC through contractual changes have emerged, ^6^ but it remains unclear how to achieve this within the current general practice framework.

Although RCC has demonstrated significant benefits, its impact varies across patient groups.^7^ For example, for patients with chronic conditions, such as diabetes or hypertension, greater continuity of care has been associated with reduced hospitalisation and mortality rates.^8^ However, it is unclear whether these benefits are similar for patients with other long-term conditions. Additionally, patients with cancer receiving RCC are more likely to be able to understand and manage their condition, experience greater feelings of control, and have higher overall well-being^9^. But there is a lack of clarity regarding the benefits of maintaining such RCC after cancer remission and recovery. A recent study showed that the productivity benefit of continuity of care seems to be greater for older patients, those with multiple chronic conditions, and individuals with mental health conditions. ^10^ However, the evidence on the differential benefits of continuity across various patient groups has not yet been synthesised.^11^

RCC may also enhance patient safety by reducing diagnostic delays, particularly for complex cases. ^12^However, while RCC fosters trust and holistic care, there are potential downsides, including diagnostic bias, patient collusion, and increased GP workload, which may contribute to clinician burnout.^13, 14, 15^

Evidence suggests that RCC is less accessible to patients from non-white ethnic backgrounds and socioeconomically deprived communities, despite their higher preference for continuity. ^16,17^ Older individuals also experience reduced RCC, highlighting systemic inequalities in access. ^18^ The implementation of RCC can be more challenging in practices situated in areas of high deprivation. This is because general practice in deprived areas is underfunded and under-doctored relative to need. ^19^ Evidence suggests a negative association between larger practice sizes and continuity, ^20^ with single-handed practices having higher patient-reported access, continuity and overall satisfaction, while higher levels of practice deprivation are associated with lower RCC. ^21^

There is an urgent need to identify effective care models that balance RCC with the realities of modern general practice. Understanding which patients benefit most, under what circumstances, and how RCC can be implemented in a multidisciplinary setting is crucial for informing policy and practice. By addressing these gaps, RCC can be optimised to improve patient outcomes, reduce health inequalities, and support the sustainability of primary care.

## Methods Research question

Implementing relational continuity in general practice - who needs it, when, to what extent, how and why?

### Aims

We aim to understand:

1. The underlying mechanisms through which RCC and its benefits are achieved
2. The populations of patients and clinicians for which RCC should be prioritised
3. The contextual conditions that constrain or facilitate RCC
4. The care models and interventions that are likely to lead to optimal implementation of RCC

### Objectives

To address our research’s aims, we will:

- Conduct a realist review of the UK and relevant international literature of RCC in general practice to develop an evidence-based programme theory of its optimal implementation
- Use the knowledge contained within the programme theory to develop relevant outputs for knowledge users and recommendations for best practice

### Approach

A realist review methodology will be used in this study. Realist reviews are particularly suited for the study of phenomena, such as RCC, which are inherently complex and multifaceted.^22^ Realist reviews aim to unpack the dynamic interactions between contexts, mechanisms, and outcomes of interest to explain causation within interventions or phenomena.^23^

### Engaging with key policy, practice and research stakeholders and with patients and public representatives

Stakeholder and Patient and Public (PPI) engagement is essential in this project. It will help us define the review’s focus and provide additional insights into the challenges and opportunities of optimally implementing RCC in general practice. It will also enable additional material (local knowledge and unpublished materials) to be identified. We will discuss emerging findings and ‘sense-check’ and further refine recommendations. This will help ensure the appropriateness of our outputs and the effectiveness of our dissemination strategy. Further, our engagement with stakeholders will leverage support for the ultimate implementation of recommendations and help to identify barriers to implementing positive changes.

We have two PPI co-applicants who have already contributed to this protocol. The PPI group participants have been purposefully selected to ensure diversity in terms of lived experience and representation of populations that have been underserved by research. The stakeholder group includes individuals who are representatives of providers, commissioners, public policy-makers and academics.

Key steps in the review will include the following:

### Step 1: Locate existing theories

The goal of this step is to identify underlying programme theories for RCC in general practice— that is, theories that explain how RCC works, in what ways, and with what consequences.

Through our early discussions and review of the literature so far, we have begun to identify the possible contexts, mechanisms, and outcomes that underlie RCC (see Figure 1). We have based this initial theory on agency theory, which proposes that the value of continuity lies in reducing agency loss by decreasing information asymmetry and increasing goal alignment between a clinician and a patient. ^24^ Continuity can help reduce information asymmetry about the patient’s health condition, needs, values, and preferences, ensuring that the clinician can tailor clinical care to the patient’s specific needs. Continuity can also assist with goal alignment, as it can be viewed as a way of forming a bond that may counteract incentives to diminish the clinician’s intention to act as the perfect agent. ^24^ Recent work by Sidaway-Lee et al^25^ attempted to summarise various factors that link continuity to patient outcomes, and we have integrated some of these factors into our initial programme theory.

**Figure 1.**
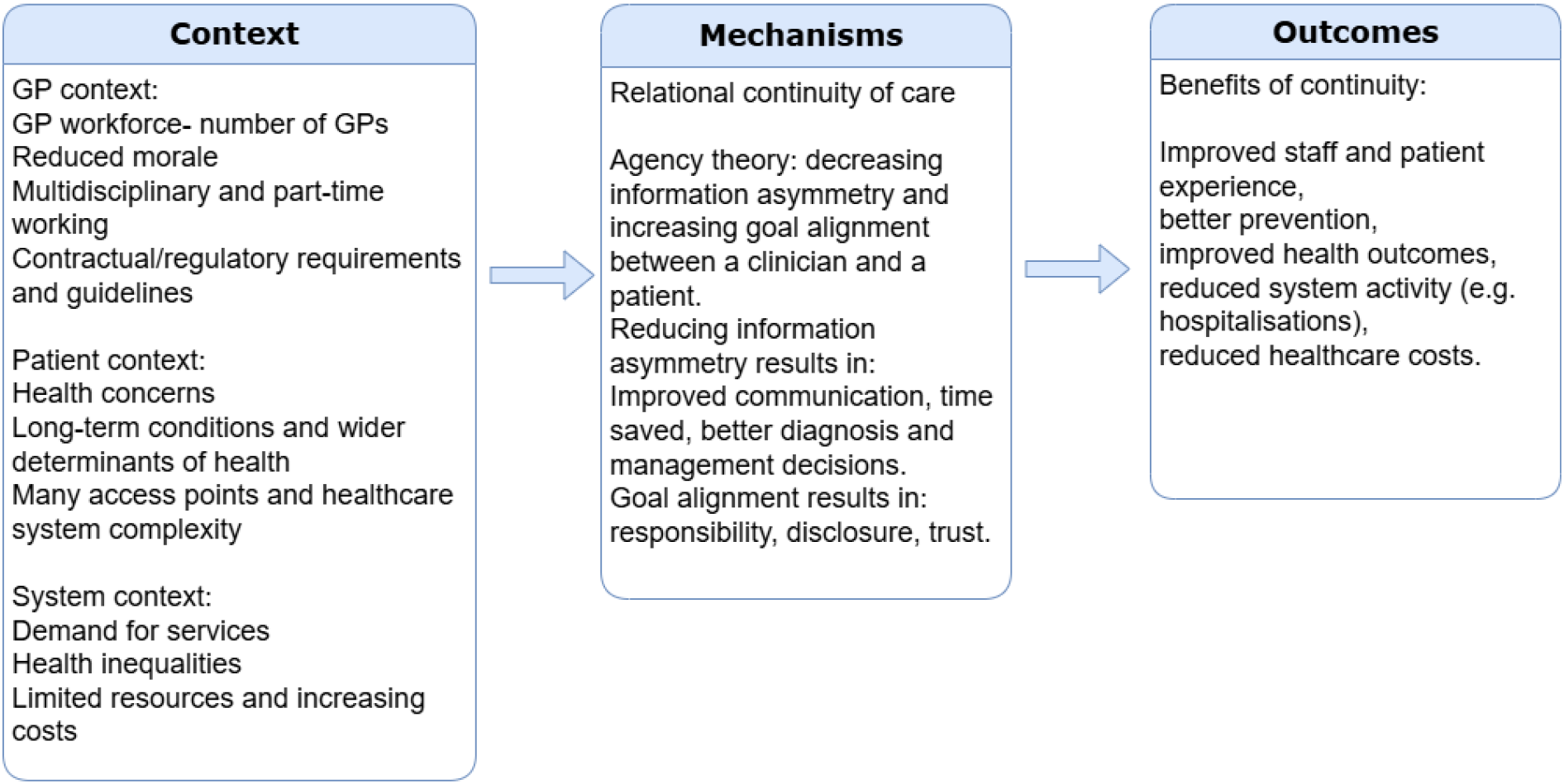
Initial Programme Theory with unconfigured possible contexts, mechanisms and outcomes of interest

This initial programme theory will be further developed in this first step of the review. We will employ a systematic method, such as the APEASE criteria^26^, to help identify the most important mechanisms within the programme theories that trigger outcomes during our meetings with stakeholders and patient and public involvement (PPI) groups.

The research team will then conduct a preliminary exploratory literature search to establish the potential scale of the evidence base, the parameters of the review and the scope. The search will apply the following search terms: continuity AND (relational OR interpersonal) AND general practice AND (interventions OR implementation OR evaluation) to MEDLINE (Ovid), Web of Science (Social Science Citation Index), Google Scholar, and the NHS Knowledge and Library Hub. Findings will inform the development of an initial programme theory or theories, which will be shared with the advisory groups for feedback, modification, and refinement.

### Step 2: Searching for evidence

The team will develop a more comprehensive search strategy, based on outputs from the previous stage. The search will be informed by the initial programme theory and by the context-intervention-mechanism-outcome framework for searching:

- Context – General practice within the UK and internationally.
- Intervention – Any intervention to facilitate RCC, including targeted (if applicable) patient populations, and any reports of experiences with such interventions by patients, their carers, and GP staff.
- Mechanism – to be informed by the preliminary literature search and discussion with the stakeholder group.
- Outcome – Improved RCC, with impacts on patient and staff experience, patient outcomes, GP workforce, service processes, and costs.

Secondary evidence will be identified from the following sources: 1) Databases - MEDLINE (OVID), EMBASE (OVID), AMED (OVID), Cochrane, CINAHLPlus (EBSCO), Academic Search

Complete (EBSCO), Web of Science (SCI, SSCI, CPCI-S, CPCI-SSH), Applied Social Sciences Index and Abstracts (ASSIA) (ProQuest), Epistemonikos, Grey literature - Health Management Information Consortium (HMIC) (OVID), ETHOS (British Library Electronic Thesis Online), Networked Digital Library of Theses and Dissertations (NDLTD) and; 2) Nuffield Trust, Kings Fund, The Health Foundation, NHS Knowledge and Library Hub. Where needed, additional searches will be developed iteratively and purposively, to enable the continuing identification of literature to inform development and testing (confirming, refuting or refining) of the programme theory.

#### Additional searching

An important process in realist reviews is searching for additional data to inform programme theory development. In other words, more searches will be undertaken if we find that we require more data to develop and test certain sub-sections of the programme theory.

### Step 3: Article selection

The results from the searching in Step 2 will be de-duplicated. There will be two stages of screening: 1) title and abstract screening; 2) full text screening.

Records will be screened by one reviewer, and at each stage, a 10% random sample will be independently screened to check for systematic errors. Disagreements will be resolved via discussion, and the involvement of a third team member if necessary. Full-text documents will be assessed for relevance, richness and rigour.

Relevance will consider whether an article aligns with the topic and supports theory development. Richness will reflect the depth of data informing Context, Mechanisms, and/or Outcomes (CMOs) within the included documents.

Rigour will follow Wong’s approach, evaluating both data sources (where needed) and the plausibility and coherence of the theoretical explanations developed – i.e. the programme theories. We will assess the plausibility and coherence of the programme theories using three criteria: consilience (whether they can explain the widest range of data), simplicity (avoiding unnecessary complexity), and analogy (alignment with existing knowledge).^27^

Where necessary, individual documents will undergo quality appraisal, particularly if they provide key evidence for a programme theory. All appraisal decisions will be transparently documented and used to report the confidence in the strength of any knowledge claims made.

### Step 4: Extracting and organising data

Data from all relevant full-text documents will be extracted using a suitably designed and piloted systematic data collection process. Key characteristics of each included document will be extracted into an Excel spreadsheet, and the full text of the documents will be uploaded to NVivo qualitative data analysis software, allowing relevant data to be organised and coded.

Coding will involve extracting relevant sections of text from included documents according to how this data can contribute to refining programme theory. This coding will be inductive (codes created to categorise data reported in included studies), deductive (codes created in advance of data extraction and analysis as informed by the initial programme theory) and retroductive (codes created based on inferences as to what may be functioning as mechanisms).

### Step 5: Evidence synthesis and drawing conclusions

Data analysis will be conducted in NVivo, and coding will use inductive, deductive and retroductive approaches to support interpretation and development of context-mechanism-outcome-configurations (CMOCs) and the programme theory or theories. We will use a series of questions to guide our realist analysis of the data from the included articles. In brief, to operationalise the realist logic of analysis, we will ask the following questions^28^:

- Interpretation of meaning: Do the documents provide data that can be interpreted as functioning as context, mechanism, or outcome?
- Interpretations and judgements about CMOCs: What is the CMOC for the data that has been interpreted as functioning as context, mechanism, or outcome?
- Interpretations and judgements about programme theory: How does this CMO relate to the initial programme theory?

Data synthesis will involve team discussions to assess the integrity of programme theories, evaluate supporting evidence, compare theories across settings, and align findings with practitioner and patient experiences. Our goal is to develop a detailed, context-specific programme theory explaining how to implement RCC in NHS general practice.

Stakeholders and PPI advisory group members will review and refine the theory through meetings and workshops, offering content expertise and real-world validation. Their feedback will ensure alignment with practical experiences, leading to a well-informed, consensus-driven final programme theory.

We will utilise data from our review, as well as feedback and advice received from workshops with our stakeholder and PPI groups, to inform the development of practical recommendations on the optimal implementation of RCC and identify evidence gaps and future research directions.

## Data Availability

This is a study protocol and there is no data available yet.

## Ethics and dissemination

### Dissemination

Our findings will be used to develop evidence-based, user-friendly, practical recommendations for the prioritisation and optimal implementation of RCC. We will collaborate with our stakeholder group, PPI members, and an artist/infographics expert to jointly develop outputs to reach different audiences, including: GP practices, policy makers, commissioners, academics, patients and the public. The study findings will be shared via conference abstracts and presentations, at least one peer-reviewed publication in a high-impact journal, a plain English summary, two end-of-study events, a series of social media infographics and a short video.

### Ethics

Ethical review is not required for this evidence synthesis as only secondary data sources will be used (QME24.1030).

## Funding statement

This project is funded by the National Institute for Health Research (NIHR) Health and Social Care Delivery Research (HSDR) (NIHR165200).

## Disclaimer

The views expressed are those of the authors and not necessarily those of the NIHR or the Department of Health and Social Care.

## Competing interests statement

VTB, KM, SP, ST and GW are practising NHS General Practitioners.

## Patient and public involvement

Patients and/or the public were involved in the design of this research protocol. Refer to the Methods section for further details. OR and SB are public co-applicants.

## Notes

### Clinical Protocols

https://doi.org/10.17605/OSF.IO/8K5XH

## References

1 Haggerty JL, Reid RJ, Freeman GK, et al. Continuity of care: a multidisciplinary review. BMJ. 2003 Nov 22;327(7425):1219–21. doi: 10.1136/bmj.327.7425.1219.

2 Sandvik H, et al. Continuity in general practice as predictor of mortality, acute hospitalisation, and use of out-of-hours care: a registry-based observational study in Norway. Br J Gen Pract. 2021. doi:10.3399/BJGP.2021.0340

3 Bazemore A, et al. The Impact of Interpersonal Continuity of Primary Care on Health Care Costs and Use: A Critical Review. Ann Fam Med. 2023 May-Jun;21(3):274–279. doi: 10.1370/afm.2961

4 Baker R, et al. (2003) Exploration of the relationship between continuity, trust in regular doctors and patient satisfaction with consultations with family doctors, Scandinavian Journal of Primary Health Care, 21:1, 27–32, doi: 10.1080/0283430310000528

5 Beech J., Opie-Martin S., Mendelsohn E., Callan C., Keith J., Fisher R. General practice data dashboard. Monitoring data on general practice. The Health Foundation. Available at: https://www.health.org.uk/general-practice-data-dashboard (xAccessed 25 July 2024)

6 Iacobucci G. Continuity of care should be “essential requirement” in GP contract, says watchdog BMJ 2023; 383 :p2839 doi:10.1136/bmj.p2839

7 Baker R, Freeman GK, Haggerty JL, et al. Primary medical care continuity and patient mortality: a systematic review. Br J Gen Pract. 2020 Aug 27;70(698):e600–e611. doi: 10.3399/bjgp20×712289

8 Chan KS, Wan EY, Chin WY, et al. Effects of continuity of care on health outcomes among patients with diabetes mellitus and/or hypertension: a systematic review. BMC Fam Pract. 2021 Jul 3;22(1):145. doi: 10.1186/s12875-021-01493-x.

9 Bulsara C, Ward AM, Joske D. Patient perceptions of the GP role in cancer management. Aust Fam Physician. 2005;34(4):299–300, 2

10 Kajaria-Montag, H., Freeman, M., Scholtes, S., Continuity of Care Increases Physician Productivity in Primary Care (June 05, 2023). INSEAD Working Paper No. 2023/23/TOM, 10.2139/ssrn.3868231

11 Fox MN, Dickson JM, et al. Delivering relational continuity of care in UK general practice: a scoping review. BJGP Open. 2024 Jun 26:BJGPO.2024.0041. doi: 10.3399/BJGPO.2024.0041.

12 Iacobucci G. Continuity of care should be “essential requirement” in GP contract, says watchdog BMJ 2023; 383 :p2839 doi:10.1136/bmj.p2839

13 Ferguson J, Stringer G, Walshe K, Allen T, Grigoroglou C, Ashcroft DM, Kontopantelis E. Locum doctor working and quality and safety: a qualitative study in English primary and secondary care. BMJ Qual Saf. 2024 May 17;33(6):354–362. doi: 10.1136/bmjqs-2023-016699.

14 Berne E (1964) Games people play. The basic handbook of transactional analysis (Ballantine Books, New York, NY

15 Wiltshire A. Improving continuity of care in general practice: four lessons from the frontline. The Health Foundation. 2019. Available at: https://www.health.org.uk/news-and-comment/blogs/improving-continuity-of-care-in-general-practice-four-lessons-from-the (xAccessed 25 March 2024)

16 Aboulghate A, Abel G, Elliott MN, et al. Do English patients want continuity of care, and do they receive it? Br J Gen Pract 2012; doi: 10.3399/bjgp12×653624

17 Stafford M, Bécares L, Hayanga B, Ashworth M, Fisher R. Continuity of care in diverse ethnic groups: a general practice record study in England. Br J Gen Pract. 2023 Mar 30;73(729):e257–e266. doi: 10.3399/BJGP.2022.0271.

18 Saunders CL, Flynn S, Massou E, et al. Sociodemographic inequalities in patients’ experiences of primary care: an analysis of the General Practice Patient Survey in England between 2011 and 2017. J Health Serv Res Policy. 2021 Jul;26(3):198–207. doi: 10.1177/1355819620986814.

19 Fisher R, Dunn P, Asaria M, Thorlby R. Briefing: Level or not? Comparing general practice in areas of high and low socioeconomic deprivation in England. 2020. Available at: https://www.health.org.uk/publications/reports/level-or-not (accessed 14 Oct 2022).

20 Forbes LJL, Forbes H, Sutton M, et al. Changes in patient experience associated with growth and collaboration in general practice: observational study using data from the UK GP Patient Survey. Br J Gen Pract 2020; doi: 10.3399/bjgp20×713429.

21 Holdroyd I, Chadwick W, Harvey-Sullivan A, Bartholomew T, Massou E, Tzortziou Brown V, Ford J. Single-handed versus multiple-handed general practices: A cross-sectional study of quality outcomes in England. J Health Serv Res Policy. 2024 Jul;29(3):201–209. doi: 10.1177/13558196231218830

22 Pawson R, et al. Realist review-a new method of systematic review designed for complex policy interventions. J Health Serv Res Policy. 2005 Jul;10 Suppl 1:21–34. doi: 10.1258/1355819054308530

23 Wong G. Realist reviews in health policy and systems research. In: Langlois ÉV, Daniels K, Akl EA, editors. Evidence Synthesis for Health Policy and Systems: A Methods Guide. Geneva: World Health Organization; 2018 Oct 8. Methods Commentary. Available from: https://www.ncbi.nlm.nih.gov/books/NBK569577/ (xAccessed 28 July 2024)

24 Donaldson MS. Continuity of care: a reconceptualization. Med Care Res Rev. 2001 Sep;58(3):255–90. doi: 10.1177/107755870105800301

25 Sidaway-Lee K, et al. What mechanisms could link GP relational continuity to patient outcomes? Br J Gen Pract. 2021 May 27;71(707):278–281. doi: 10.3399/bjgp21×716093.

26 Michie, S., Atkins, L., & West, R. (2014). The Behaviour Change Wheel: A Guide to Designing Interventions. Great Britain: Silverback Publishing

27 Wong G. In: Emmel N, Greenhalgh J, Manzano A, Monaghan M, Dalkin S, editors. Data gathering for realist reviews: Looking for needles in haystacks. Doing Realist Research. London: Sage, 2018

28 Wong, G., et al. Interventions to improve antimicrobial prescribing of doctors in training: The IMPACT (IMProving Antimicrobial presCribing of doctors in Training) realist review. 2015. BMJ Open, 5(10), 1–8. 10.1136/bmjopen-2015-009059

